# Genetics of cardiovascular outcomes in individuals with chronic kidney disease: the Chronic Renal Insufficiency Cohort (CRIC) study

**DOI:** 10.1101/2025.02.19.25322572

**Authors:** Jia Wen, Bridget M Lin, Quan Sun, Min-Zhi Jiang, Greg Linchangco, Gang Li, Ruochen Chen, Alan S. Go, Tyne W. Miller-Fleming, Megan M. Shuey, Debbie L. Cohen, Panduranga S. Rao, Mahboob Rahman, Nancy J. Cox, James P. Lash, Wyliena Guan, Daniel C. Posner, Qin Hui, Serena C. Houghton, Adriana M. Hung, Kelly Cho, Peter W.F. Wilson, Haibo Zhou, Yan V. Sun, Yun Li, Nora Franceschini, Million Veteran Program and the CRIC Study Investigators

**Author notes:** Corresponding authors: Nora Franceschini, MD, MPH, Departments of Epidemiology and Biostatistics Yun Li, PhD, University of North Carolina, Chapel Hill, NC. Shared authors (Jia Wen, Bridget M Lin, Yun Li, Nora Franceschini).

## Abstract

Genome-wide association studies (GWAS) identified multiple loci for cardiovascular disease, but their relevance to individuals with chronic kidney disease (CKD), who are at higher risk of cardiovascular disease, is unknown. In this study, we performed GWAS analyses of coronary heart disease (CHD) or all-cause stroke in African (AFR) and European (EUR) American participants with CKD of the Chronic Renal Insufficiency Cohort (CRIC). Mixed-effect logistic regression models were race-stratified and adjusted for age, sex, site of recruitment, estimated glomerular filtration rate (eGFR), and principal components, followed by meta-analysis. We attempted replication in participants from two biobanks with biomarker or ICD-10 (International Classification of Diseases, 10th Revision) diagnostic codes for CKD. We assessed the association of single nucleotide variants (SNVs) at known CHD and stroke loci in CRIC and tested the genetic correlation among CRIC, a biobank-based cohort and published GWAS of cardiovascular disease. Among 3,588 CRIC participants, 1,203 had CHD and 535 had all-cause stroke. We identified six SNVs across three loci (*LINC02744*, *AZIN1- AS1*, and *ATP6V0A4*) associated with all-cause stroke, and two intronic SNVs at the *PPARG* locus associated with CHD. However, SNV associations were not significant in replication studies. Published SNVs for CHD or stroke were not associated with cardiovascular outcomes in CRIC. When testing the genetic correlations between published GWAS and CRIC GWAS, they were significant for CHD (genetic correlations (rg) range of 0.39 to 0.51, *p-value<*0.007). These findings suggest some differences in the genetic architecture of CHD and stroke among individuals with CKD compared to those from the general population, although large numbers of CKD participants are needed to assess if findings are related to participant selection and CKD severity, or non-traditional risk factors in people with CKD.

## Introduction

Cardiovascular disease is the leading cause of death worldwide, with an estimated 17 million annual deaths, and is a major contributor to disability^1^. Individuals with chronic kidney disease (CKD) are at high risk for atherosclerotic and non-atherosclerotic cardiovascular events^2,3^, as well as developing cardiovascular disease at younger ages^2,4,5^. Acute myocardial infarction (MI) is four times more common in patients with CKD than without CKD, while stroke is twice as common^6,7^. The burden of cardiovascular disease increases with worsening estimated glomerular filtration rate (eGFR) and advanced CKD stages^8^. CKD occurs in 10% of U.S. adults and approximately 15% of Medicare-eligible individuals^3^. Therefore, individuals with CKD contribute to a substantial burden of cardiovascular disease in the U.S.

The underlying mechanisms for increased cardiovascular disease in individuals with CKD are likely multifactorial. Several non-traditional risk factors for cardiovascular disease have been identified in individuals with CKD, including inflammation, abnormalities in calcium-phosphorus metabolism, anemia and oxidative stress^2,9^. However, the genetic susceptibility to cardiovascular disease in individuals with CKD has been less explored. Prior genome-wide association studies (GWAS) have identified several loci for coronary heart disease (CHD), stroke, and other cardiovascular outcomes^10–12^, but the relevance of these findings to individuals with CKD is unknown, given potential differences in disease susceptibility and risk factors.

To uncover the genetic susceptibility to cardiovascular disease in individuals with CKD compared to those in the general population, we performed GWAS of CHD and all-cause stroke in participants of the Chronic Renal Insufficiency Cohort (CRIC)^13,14^. We then compared CRIC findings to published studies of individuals not selected for CKD and to GWAS of Million Veterans Program (MVP) participants with ICD-10 diagnostics codes for CKD. CRIC recruited individuals with an eGFR between 20 to 70 mL/min/1.73m^2^ from U.S. clinical centers. CRIC participants have a high burden of cardiovascular disease, and several non-genetic risk factors for cardiovascular disease have been identified in this cohort^13,14^.

## Results

The mean age for CRIC participants was 57.79 years, 55% were men, and the mean eGFR was 52.31 ml/min/1.73 m^2^ (**Table 1**). The total number of individuals by inferred ancestry and number of events for each outcome are shown in **Table S1** for the primary analyses and analyses restricted to incident events.

**Table 1.** Characteristics of participants in CRIC.

### GWAS findings in the CRIC study

**Table 2** shows main findings for meta-analysis of CHD and all-cause stroke. Ancestry-specific results for index variants are shown in **Table S2.** Given the AFR and EUR results were driven by low-frequency variants, all follow-up analyses were based on meta-analysis results. Manhattan plots and QQ plots for meta-analyses of CHD and all-cause stroke are shown in **Figure 1 and Figure 2** respectively. There was little evidence for inflation (λ =1.001 for CHD and λ =0.998 for all-cause stroke). We identified 6 SNVs across three loci (*LINC02744*, *AZIN1-AS1*, and *ATP6V0A4)* associated with all-cause stroke, and 2 SNVs at the *PPARG* locus associated with CHD (rs13088728, rs7620165, MAF > 3%, intronic to *PPARG*) (**Table 2 and Table S2**). Findings from meta-analyses for time-to-event incident events (**Table S3)** were not the same with primary analysis. Detailed results from time-to-events analyses are described in supplemental material.

**Figure 1.**
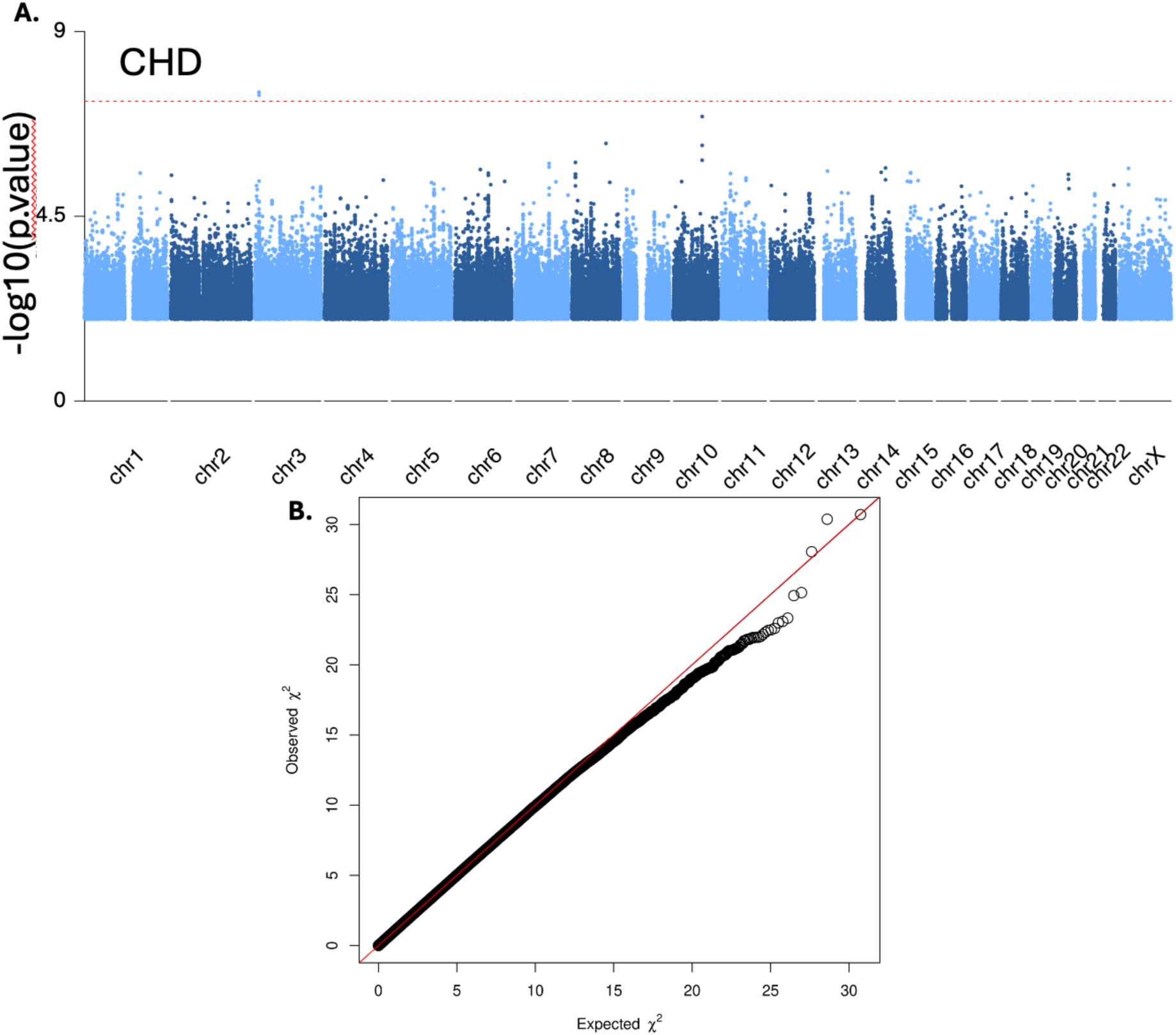
Meta-analyzed results for CHD in CRIC. **A**. Manhattan plots for CHD in CRIC. X- axis lists the chromosome position for each variant and Y-axis shows the -log10(*p*.value) for associations. **B**. Quantile-quantile (QQ) plot of GWAS results for CHD in CRIC.

**Figure 2.**
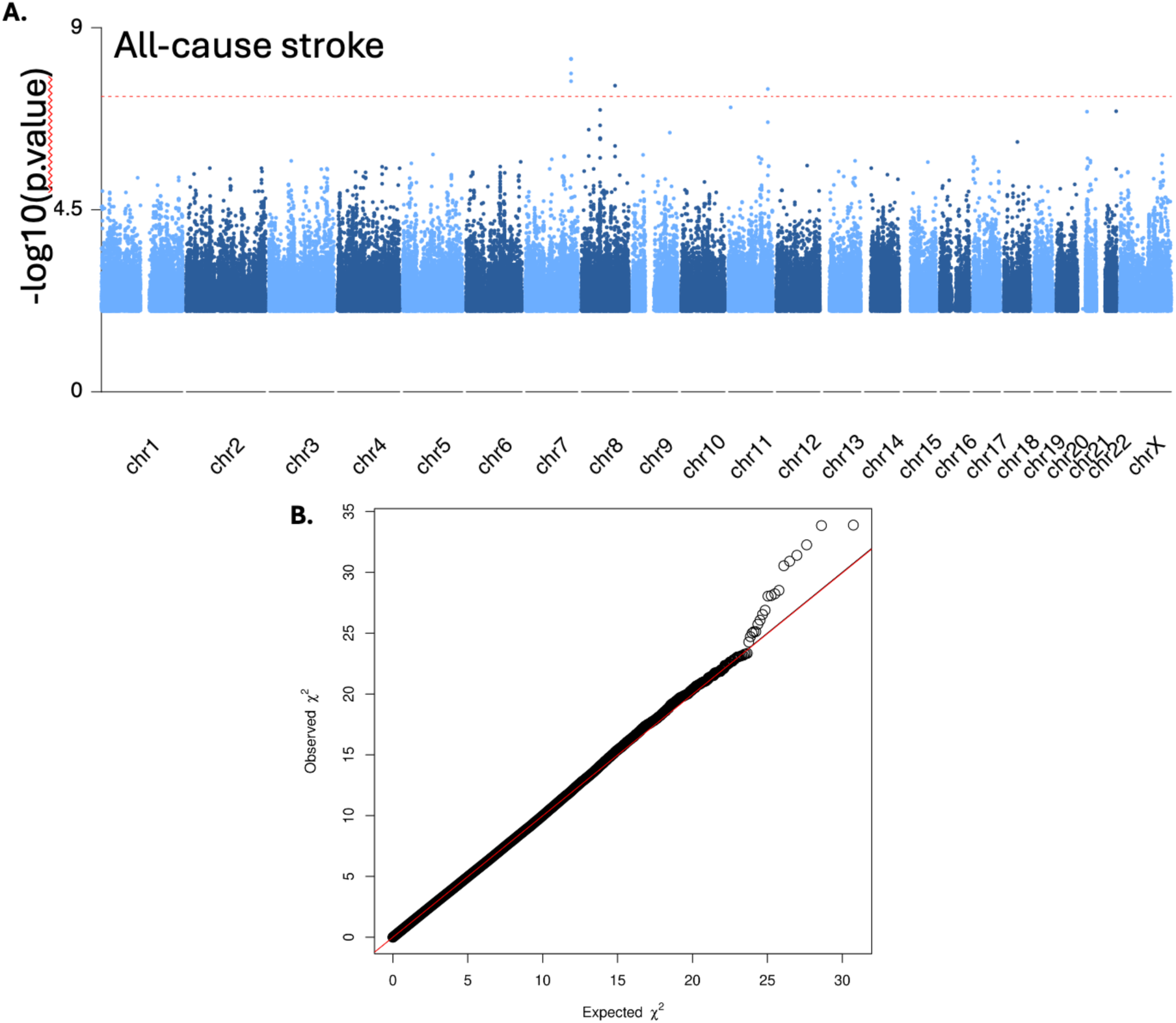
Meta-analyzed results for all-cause stroke in CRIC. **A**. Manhattan plots for all-cause stroke in CRIC. X-axis lists the chromosome position for each variant and Y-axis shows the - log10(*p*.value) for associations. **B**. Quantile-quantile (QQ) plot of GWAS results for all-cause stroke in CRIC.

**Table 2.** Significant GWAS AFR/EUR CRIC meta-analysis results for CHD and all-cause stroke.

### Comparison of our findings with published GWAS findings

We used CRIC GWAS meta-analyses of CHD and stroke to assess the association of SNVs identified in published EUR or multi-population GWAS meta-analyses for CHD (198-247 loci^10,11^) and stroke (8 - 48 loci^15,16,12^) (SNVs included in **Table S5** and published GWAS in **Table S6**). The published SNVs were not associated with CHD or stroke in the CRIC GWAS meta-analysis results (**Table S7)** or when we assessed only CRIC EUR participants (**Table S8**). Not replicating literature-reported variants in CRIC may reflect distinct genetic architecture or lack of statistical power. The estimated power for detecting known variants was found to be low (<5%) in CRIC with the limited sample size. Our findings in MVP, where we had larger sample sizes and where several literature-reported CHD and stroke loci **(Table S9**) were replicated, support that the smaller sample size is a main contributor to the lack of replication. However, it is interesting to observe that the number of loci replicated in MVP was equal or less than expected (**Table S9**), likely resulting from a combination of winner’s curse (effect sizes for published variants tend to be over-estimated) and at least partially distinct genetic architecture of cardiovascular outcomes among individual with CKD versus among the general population.

### GWAS genetic correlations

To directly assess the level of shared genetic architecture, we estimated and tested the genetic correlations among MVP CKD GWAS, CRIC GWAS and published GWAS for CHD and stroke (**Table S4,** and **Figure 3**). Note that the first two are among individuals with CKD while published studies were derived from the general population without considering CKD status. The genetic correlations between MVP and CRIC were not significant for CHD and all-cause stroke (**Table S4**) which may be due to the differences in definitions of CKD in MVP (based on ICD-10 diagnostic codes for CKD) and CRIC (clinical disease). However, the genetic correlations were significant for CRIC CHD meta-analysis and published CHD EUR and multi-population GWAS^10,11^ (rg = 0.39 and 0.42 and *p*.values = 0.0025 and 0.0009, respectively) and when testing the CRIC EUR and published CHD EUR and multi-population GWASs (rg = 0.51 and 0.47, and *p*.values = 0.0068 and 0.005, respectively)^10,11^ (**Table S4,** and **Figure 3**). The MVP GWAS also showed significant genetic correlation with published EUR CHD GWAS (**Table S4,** and **Figure 3**). We were unable to estimate the genetic correlations for all-cause stroke between CRIC and the published GWAS of stroke due to the out-of-bound heritability estimation from LDSC.

**Figure 3.**
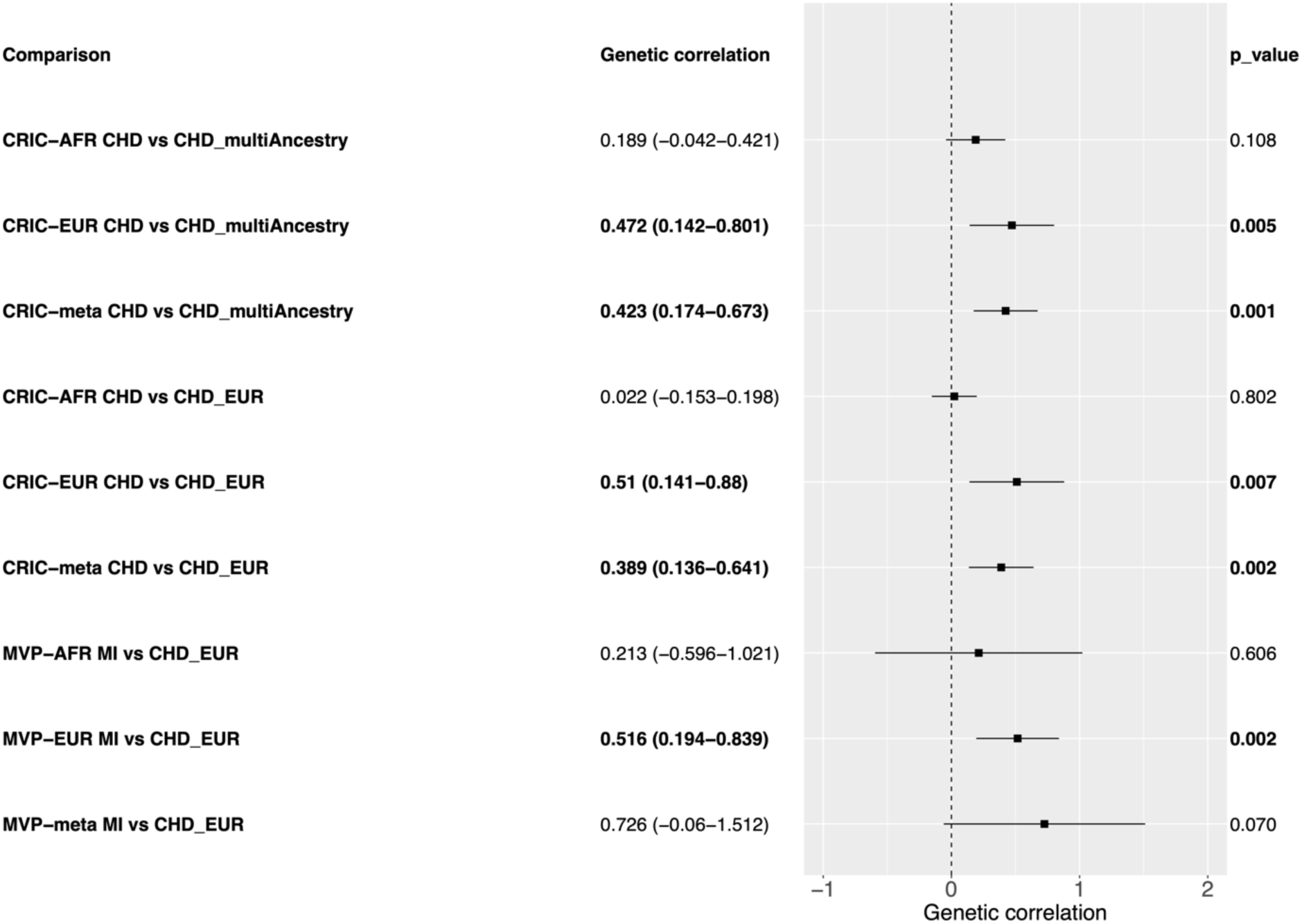
The forest plot shows the genetic correlation results. The bold font denotes the significant genetic correlation at *p*.value < 0.05. The number in the parentheses shows the confidence interval of the genetic correlation estimate. The dots denote the genetic correlation estimates and the lines denote the confidence interval.

## Discussion

This study provides some insights into the study of genetic susceptibility to cardiovascular disease in individuals with CKD, who are at high-risk for CHD and stroke. Studies of participants with clinically defined CKD, such as CRIC, have small samples and low power for discovery or replication. For example, the *PPARG* locus was associated with overall CHD in meta-analysis of EUR and AFR (**Table 2**), but the finding did not replicate. We also identified three loci for stroke, which did not replicate. Failing to replicate literature-reported loci is not entirely surprising. Interestingly, replication rate in the larger sample size MVP was equal to or lower than expected, suggesting at least partially distinct genetic architecture of cardiovascular outcomes in the general population versus individuals with CKD. To assess and quantify the extent of shared and distinct genetic determinants of cardiovascular outcomes between the studies, we performed genetic correlations among MVP, CRIC, and published GWAS for CHD and stroke. Note that published GWAS are conducted in studies without considering CKD status, while both MVP and CRIC individuals were selected for CKD, with the main difference being that CKD is defined in MVP using ICD-codes while CRIC recruited individuals from CKD clinics. Non-significant genetic correlations between MVP and CRIC results, even when we studied only EUR individuals, points to differences in the genetic architecture of CKD participants selected from a biobank and a clinical setting. However, there are also differences in demographics such as sex (e.g., larger proportion of males in MVP than CRIC) and age, which relates to differences in CKD causes (**Table 1**). Interestingly, MVP GWAS showed strong and significant genetic correlations with published GWAS of individuals not selected for CKD, suggesting larger genetic similarity to cardiovascular outcomes among these two studies. This is likely due to misclassification of CKD when using ICD-10 diagnostic codes and the larger sample of the MVP studies, rendering MVP more similar to the published studies than CRIC is. Finally, despite the small sample, we found significant genetic correlations between CRIC and published GWAS for CHD. Specially, larger genetic correlations were observed among CRIC EUR GWAS than CRIC meta GWAS when compared to either published EUR GWAS of CHD or those GWAS that included multi-populations. The published studies are large meta-analyses that likely included a small number of individuals with CKD given cardiovascular disease is prevalent among older individuals who have CHD. The results suggest some overlap in genetic factors contributing to cardiovascular disease. However, genetic correlations were not strong, and differences in findings may be related to ancestry and sample size but also mechanisms related to advanced CKD such as in individuals from the CRIC study. Individuals with advanced CKD have accelerated atherosclerosis and cardiovascular events not related to atherosclerosis such as arrhythmias. Therefore, additional genetic factors may contribute to cardiovascular disease in individuals with CKD

In summary, we identified differences in cardiovascular genetic factors among individuals with and without CKD that will need to be further assessed in larger and well-powered studies. The new insights are important to understand disease mechanisms and potential approaches for treatment of cardiovascular disease in individuals with CKD.

## Methods

### Study description

CRIC is a multicenter and multi-ethnic cohort study that enrolled 3,939 participants aged 21 to 74 years with mild-to-moderate CKD from seven U.S. clinical centers between 2003 and 2008. The study design and participant characteristics have been previously published^17,18^. Briefly, eGFR entry criteria was 20 to 70 mL/min/1.73m^2^. Individuals with severe heart failure, cirrhosis, HIV infection, polycystic kidney disease, renal cell carcinoma, those who received chronic dialysis or a kidney transplant, and those taking immunosuppressant drugs were excluded. CRIC has detailed information on demographics, medical history, and medications at baseline and subsequent annual visits in addition to physical measures obtained using standardized methods. eGFR was estimated using the race-free 2021 CKD-EPI equation^19^ The study was approved by the institutional review boards from each of the participating clinical centers, and all participants provided written informed consent.

### Cardiovascular events

Cases were defined to include both prevalent and incident events. Prevalent cases are defined at baseline visit self-report history of MI or coronary revascularization (for CHD) or history of stroke. Cardiovascular incident events were identified through annual clinical visits and bi-annual telephone interviews based on hospitalizations, outpatient tests or interventions, and adjudicated by review of medical records. Briefly, MI was defined by chest pain, electrocardiogram abnormalities, and elevated cardiac biomarkers. Stroke was determined on autopsy findings or by sudden onset of neurologic symptoms lasting >24 hours supported with imaging (demonstration of infarction in a territory where an injury or infarction would be expected to create those symptoms). Incident strokes were classified as ischemic or hemorrhagic using available information from medical records and imaging studies. Death was obtained by reports by next of kin, death certificates, hospital records, and linkage with the Social Security Death Master File. The cause of death (including sudden cardiac death vs non-cardiovascular disease) was ascertained during hospitalization through medical records.

### Genotypes, imputation methods and quality control

CRIC participants were genotyped at the Broad Institute using the Illumina HumanOmni 1-Quad Array Platform. Genotypes were called using Illumina’s BeadStudio Genotyping Module (Illumina Inc, San Diego, CA, USA). Quality control of genotyped data has been previously reported^20^. The final genotyped dataset included 918,638 variants.

The CRIC genotyped sample included 1,723 self-reported white individuals, 1,525 self-reported Black individuals, and 385 individuals missing self-reported race/ethnicity. We used HARE, a machine learning algorithm that produces a surrogate variable, to infer ancestry in these samples^21^. We excluded 29 individuals with undetermined HARE ancestry and 4 individuals with sample duplicates and 12 with sex mismatch (total n=44 excluded). Our final data based on complete phenotype and genotype data included 2,070 individuals inferred as European (EUR) ancestry and 1,518 individuals inferred African (AFR) ancestry, for a total of 3,588 individuals. Principal components (PCs) were estimated from genotypes across all samples using standard procedures^22^. Imputation was performed to the TOPMed (Trans-Omics for Precision Medicine) reference panel (freeze 5b) using minimac4 via the Michigan Imputation Server^23^. Genetic variants with imputation quality Rsq > 0.3 and minor allele frequency > 0.01 within EUR or AFR samples were considered passing quality control (QC) and were used in the GWAS analysis.

### Statistical analysis

For our primary analysis, we defined the overall CHD and all-cause stroke by combining incident (including fatal events) and prevalent cases to mirror definitions used in published GWAS that included case-control and incident event studies. We also tested the associations with incident CHD or all-cause and ischemic stroke by excluding individuals with prevalent disease in time-to-event analyses. We tested the association of each single nucleotide variant (SNV) with overall CHD and all-cause stroke using mix-effect logistic regression models implemented by Generalized linear Mixed Model Association Tests (GMMAT)^24^. The kinship matrices of genetic relatedness between pairs of individuals in AFR and EUR separately were estimated using EPACTS v3.3.0. In the mixed-effect logistic regression model, the kinship matrices were used as random effects with adjustments for age, sex, site of recruitment, baseline race-free eGFR, and top 10 genotype PCs in analyses stratified by EUR and AFR individuals. GWAS results were then combined using fixed-effect meta- analysis^25^ for a total of 16.7 M variants common to both samples tested. The genome-wide significance threshold was set to be 5x10^-8^. A locus was defined by a 1MB window from the most significant variant (index variant) in the meta-analyzed results. If the index SNV resided within a gene (exonic or intronic), the gene name was taken as the locus name; for intergenic index SNVs, the nearest gene is taken as the locus name. For each SNV identified in CRIC, we checked whether there was any overlap with significant published SNVs for CHD or stroke using a 1 MB window and linkage disequilibrium (LD) *r^2^*>0.8 (in both AFR and EUR from TOPLD^26^) with the CRIC significant SNV.

### Replication of CRIC novel GWAS findings in the UK Biobank and MVP participants with CKD

We attempted replication of new associations in the UK Biobank, using the subset of EUR participants with CKD. The UK Biobank is a prospective study of over 500,000 participants from England, Scotland, and Wales, aged 40 and 69 years at recruitment (2006-2010)^27^. Participants completed a comprehensive baseline assessment including physical measures, and clinical and phenotypic data on biomarkers is available. All participants provided written informed consent. To be comparable with our discovery study, we selected the subset of UK Biobank participants with CKD based on having an eGFR<60 ml/min/1.73 m^2^ or a urine albumin to creatinine ratio ≥300 mg/g at the baseline assessment^19^. Participants reporting cardiovascular disease before or at the baseline assessment date were included in the primary analysis results replication. Analysis was restricted to those of European ancestry (n=8,433) due to small number of non-EUR individuals with CKD. Ancestry was determined based on a combination of self-reported and k-means clustering, as described in previous publications ^28^. The UK Biobank outcomes available were CHD, and all-cause stroke obtained through algorithms developed using the UK Biobank linked Hospital Inpatient and Death Registry data^29,30^. UK Biobank participants were genotyped using two arrays (UK BiLEVE Axiom Array/UK Biobank Axiom Array). We performed quality control and imputation to TOPMed reference genomes (freeze 8). For replication, we tested the association of SNVs with a minor allele frequency (MAF) > 0.03 from CRIC meta-analyses that had an imputed Rsq > 0.3 in the UK Biobank. We used mixed-effect logistic regression implemented in GMMAT, for models adjusted for age, sex, recruitment site, baseline race-free GFR and top 10 genotype PCs. We compared the estimates for the direction of effects in addition to p-values between discovery and replication findings.

We also attempted replication in the Million Veteran Program (MVP) participants with CKD. We selected participants with CKD based on ICD-10 diagnostic codes for CKD and performed GWAS for CHD and Acute ischemic stroke in EUR and AFR participants. MVP CHD primary cardiovascular outcomes of interest included incident myocardial infarction, acute ischemic stroke, and cardiovascular disease (CVD)-related death. Incident myocardial infarction and acute ischemic stroke events in MVP were similarly defined using ICD-10 codes, ensuring consistency across the phenotype definitions. We performed GWAS for CHD and stroke using REGENIE and SNV with a MAF> 0.01. Models were adjusted by age, sex and principal components^31^. The MVP study has got approved and performed in accordance with relevant guidelines.

### Comparison of CRIC GWAS results with published GWAS and MVP study with participants selected for CKD

We assessed the evidence for association in CRIC and MVP of published SNVs using summary statistics from published GWAS meta-analyses from multi-population and European-ancestry CHD^10,11^, and stroke and subtypes^15,16,12^. We then calculated statistical power in CRIC using effect size and effect allele frequency of known variants in corresponding GWAS, to investigate the probability of replicating known variants in CRIC. We additionally used data from the MVP GWAS for CHD and stroke in EUR and AFR in individuals with CKD to estimate the number of replicated SNVs. We then tested the genetic correlation among the MVP CHD and stroke GWAS, published GWAS analyses and CRIC GWAS using Linkage Disequilibrium Score Regression (LDSC) method^32^.

### Annotation of SNVs

We annotate the SNVs using TopLD for linkage disequilibrium and functional annotation^26^. The SNVs of meta-analyzed results are intronic variants of corresponding genes and some of them reside in transcript region targeting nonsense-mediated mRNA decay.

## Supporting information

Main tables

## Acknowledgments

Funding for this project was obtained through the CRIC Study Opportunity Pool Program and NIH R01-HL163972 to NF and YL. Funding for the CRIC Study was obtained under a cooperative agreement from National Institute of Diabetes and Digestive and Kidney Diseases (*U01DK060990, U01DK060984, U01DK061022, U01DK061021, U01DK061028, U01DK060980, U01DK060963, U01DK060902 and U24DK060990*). In addition, this work was supported in part by: the Perelman School of Medicine at the University of Pennsylvania Clinical and Translational Science Award NIH/NCATS *UL1TR000003*, Johns Hopkins University *UL1 TR-000424*, University of Maryland *GCRC M01 RR-16500*, Clinical and Translational Science Collaborative of Cleveland, *UL1TR000439* from the National Center for Advancing Translational Sciences (NCATS) component of the National Institutes of Health and NIH roadmap for Medical Research, Michigan Institute for Clinical and Health Research (MICHR) *UL1TR000433*, University of Illinois at Chicago CTSA *UL1RR029879*, Tulane COBRE for Clinical and Translational Research in Cardiometabolic Diseases *P20 GM109036*, Kaiser Permanente NIH/NCRR *UCSF-CTSI UL1 RR-024131*, Department of Internal Medicine, University of New Mexico School of Medicine Albuquerque, *NM R01DK119199*. JW was partially supported by NIH T32ES007018. WG was supported by a grant from the National Institutes of Health, USA (NIH grant *T32ES007018-46*). This research is based on data from the Million Veteran Program, Office of Research and Development, Veterans Health Administration, and was supported by award # BX004821. This publication does not represent the views of the Department of Veteran Affairs or the United States Government.

## Author contributions

Conceptualization: YL, NF. MVP GWAS provider: GL, YVS. Data QC: JW, BL, GL. Study Design and analysis: JW, BL, MJ, G Linchangco, G Li, RL, YL, NF. Replication: JW, RL, TM, MS. Investigation: SY, YL, NF. Visualization: JW. Supervision: YL, NF. Writing: JW, NF. All authors review and edit the manuscript.

## Data availability statement

Data from the Chronic Renal Insufficiency Cohort Study [(V12)/https://doi.org/10.58020/6dxf-ed78] reported here are available for request at the NIDDK Central Repository (NIDDK-CR) website, Resources for Research (R4R), https://repository.niddk.nih.gov. The summary statistics for the GWAS of CHD and stroke will be deposited at the Genome Catalog. The MVP GWAS results are available from the corresponding author upon reasonable request.

## Additional information Competing interests

The authors declare no competing interests.

---

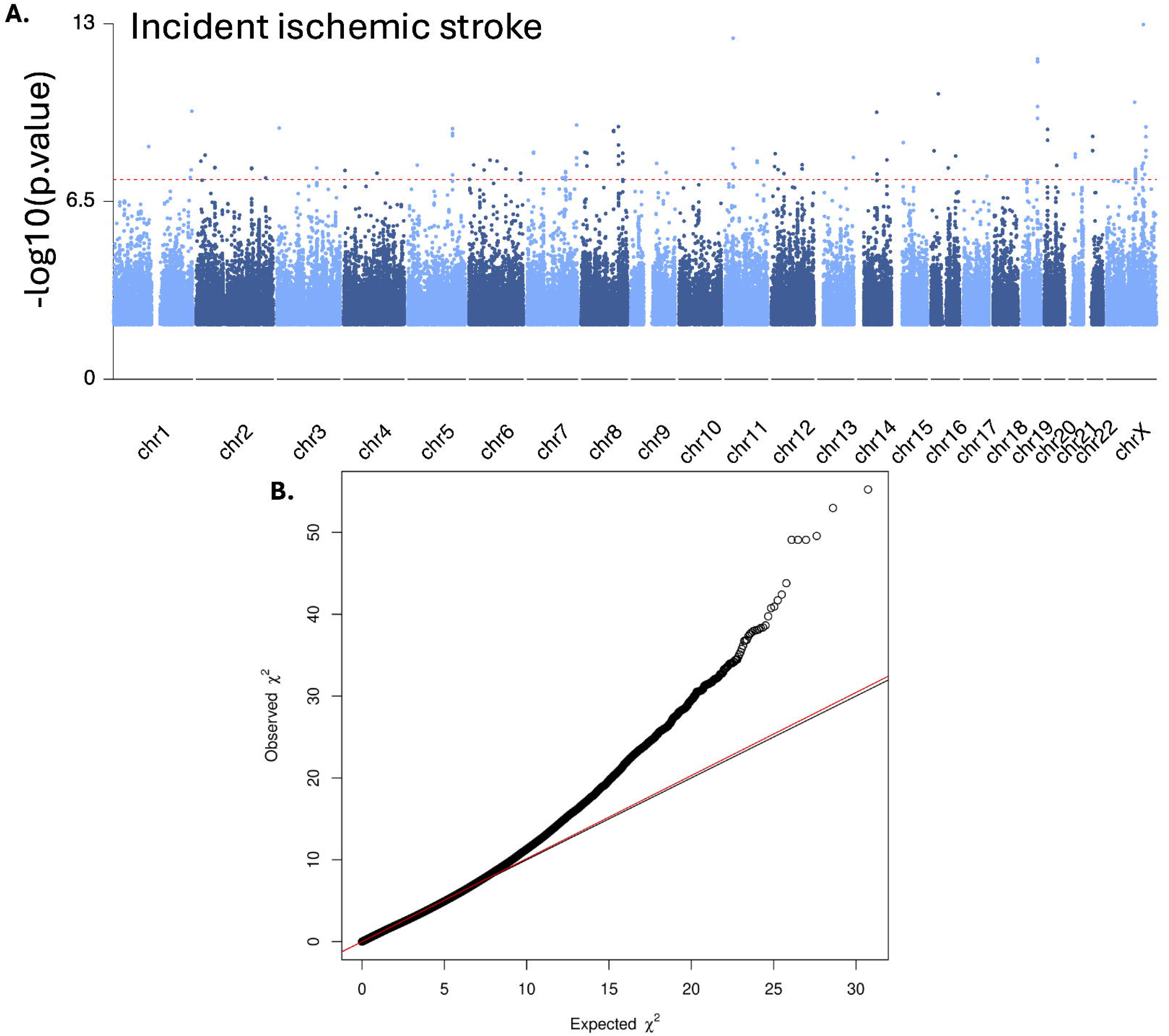

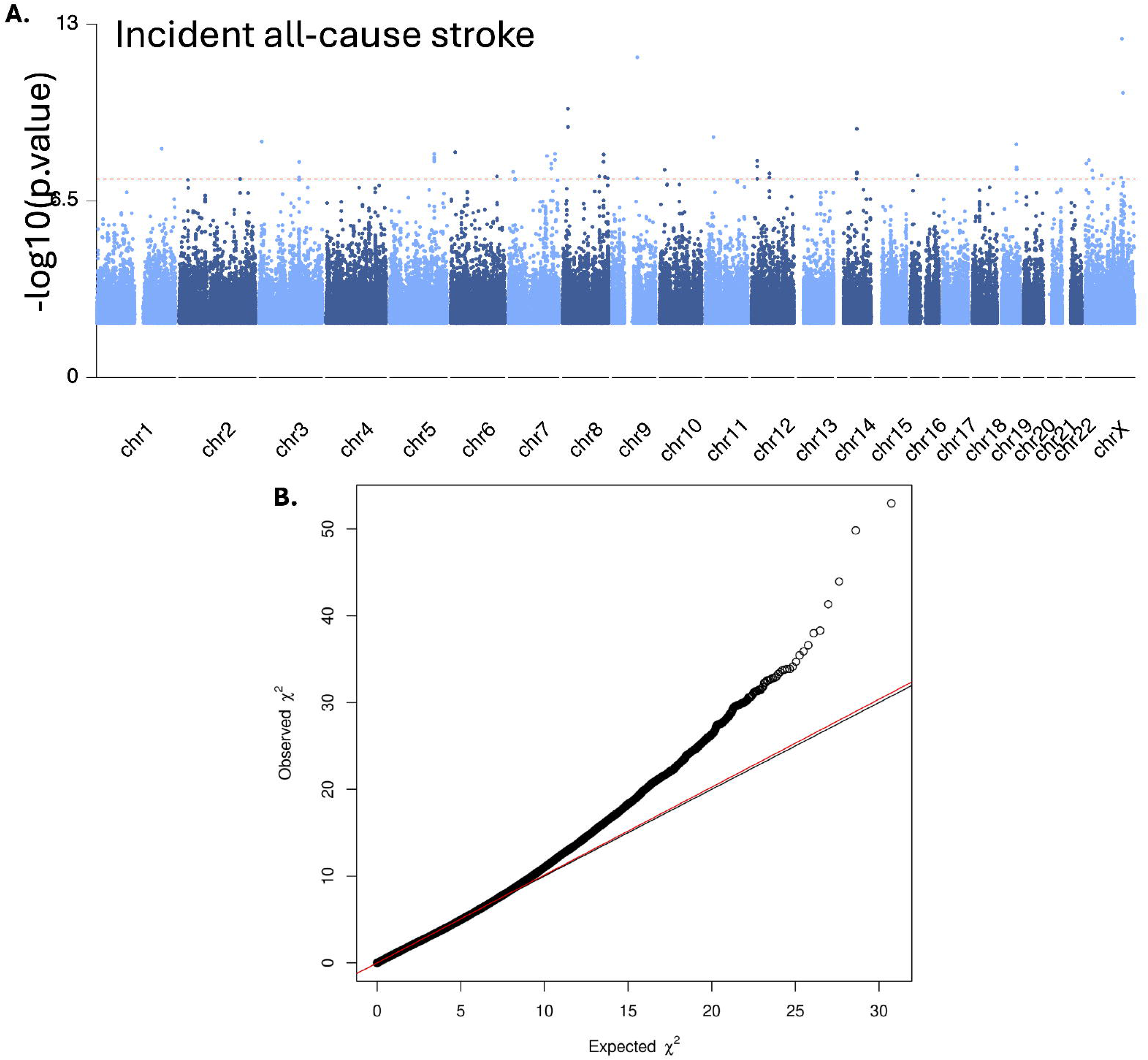

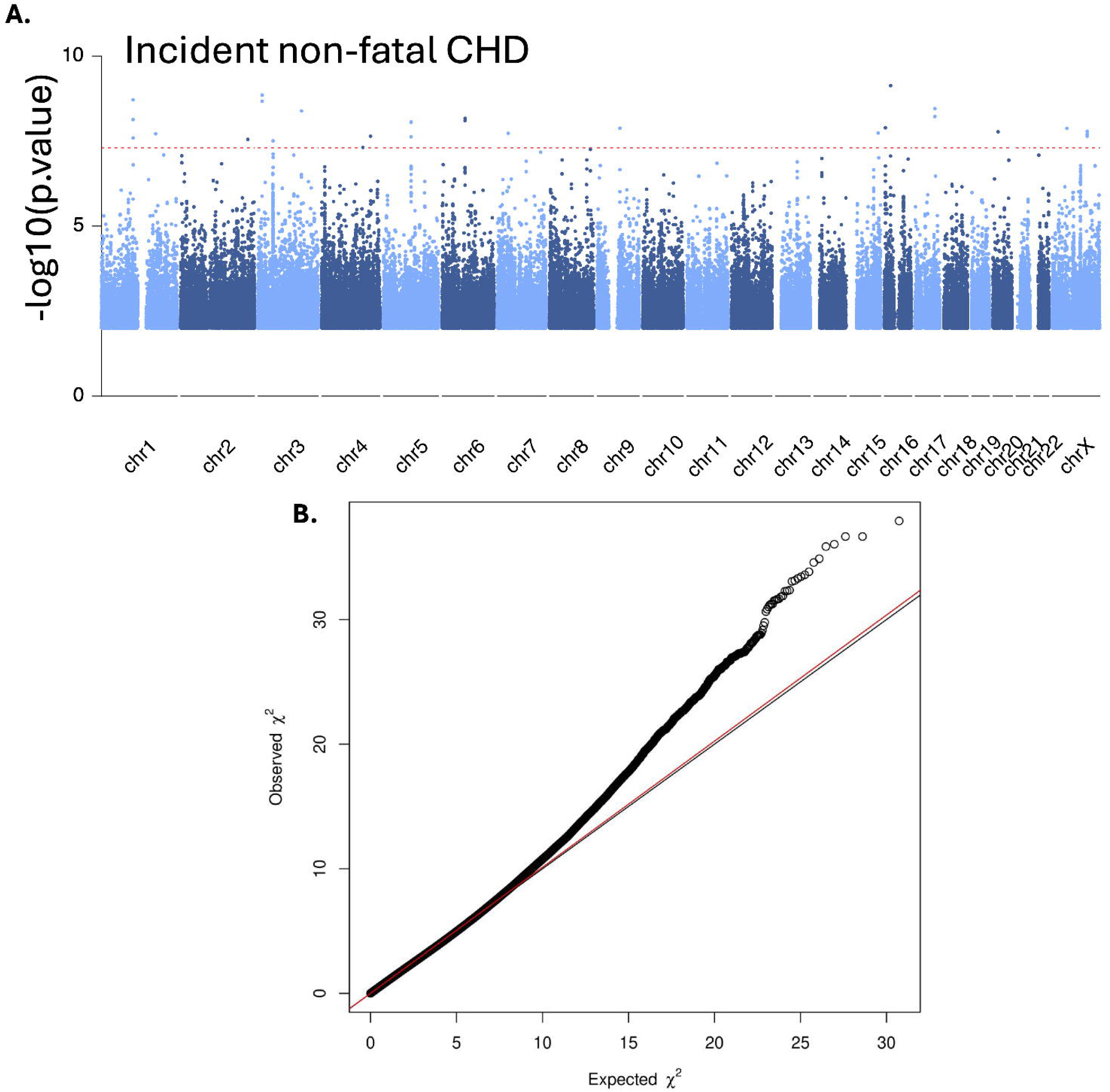

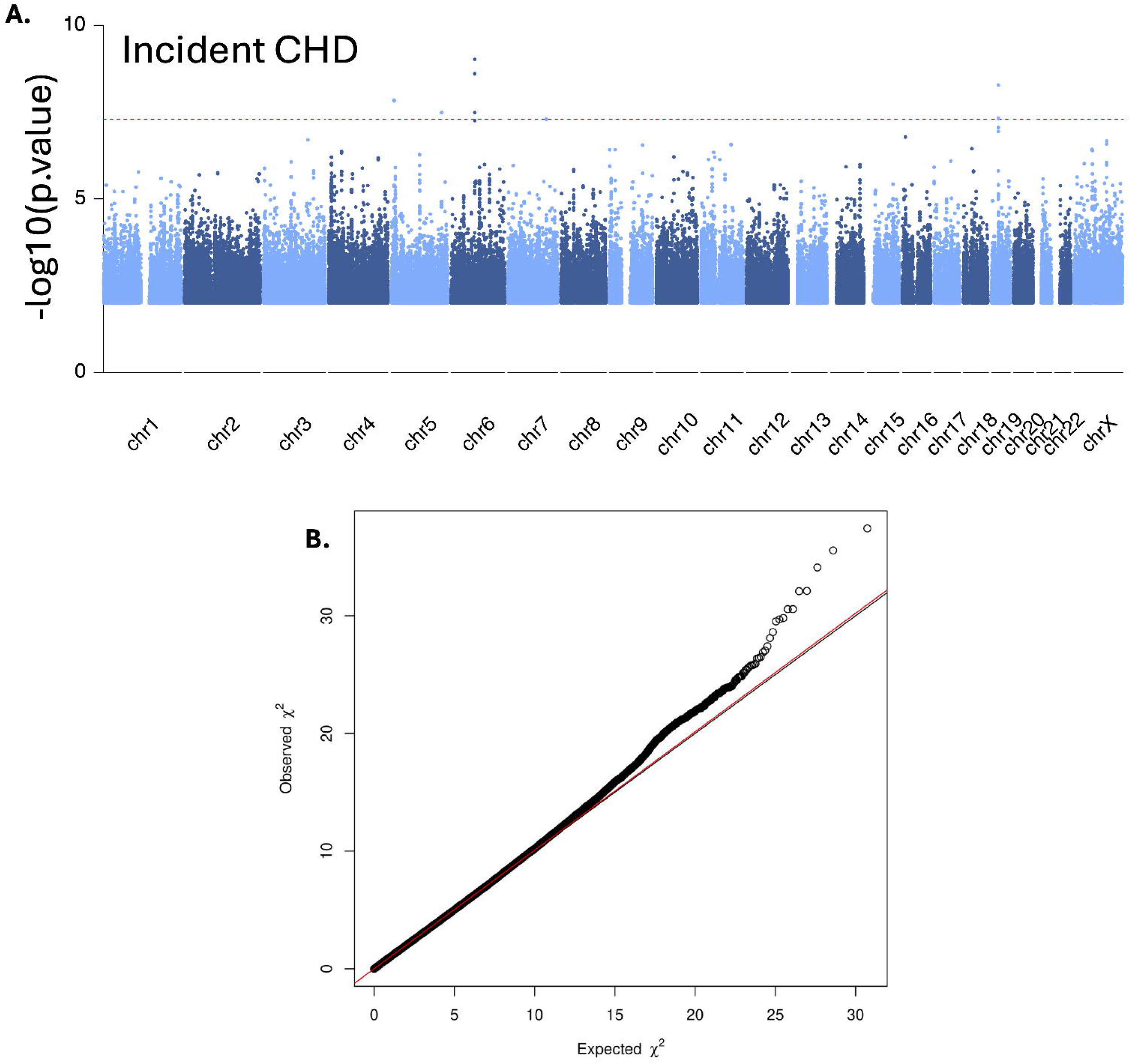

## Reference

1 Roth, G. A. et al. Global Burden of Cardiovascular Diseases and Risk Factors, 1990-2019: Update From the GBD 2019 Study. J Am Coll Cardiol 76, 2982-3021, doi:10.1016/j.jacc.2020.11.010 (2020).

2 Sarnak, M. J. et al. Chronic Kidney Disease and Coronary Artery Disease: JACC State-of-the-Art Review. J Am Coll Cardiol 74, 1823–1838, doi:10.1016/j.jacc.2019.08.1017 (2019).

3 Johansen, K. L. et al. US Renal Data System 2020 Annual Data Report: Epidemiology of Kidney Disease in the United States. Am J Kidney Dis 77, A7–A8, doi:10.1053/j.ajkd.2021.01.002 (2021).

4 Virani, S. S. et al. Heart Disease and Stroke Statistics-2020 Update: A Report From the American Heart Association. Circulation 141, e139–e596, doi:10.1161/CIR.0000000000000757 (2020).

5 Go, A. S., Chertow, G. M., Fan, D., McCulloch, C. E. & Hsu, C. Y. Chronic kidney disease and the risks of death, cardiovascular events, and hospitalization. The New England journal of medicine 351, 1296–1305, doi:10.1056/NEJMoa041031 (2004).

6 Di Angelantonio, E., Danesh, J., Eiriksdottir, G. & Gudnason, V. Renal function and risk of coronary heart disease in general populations: new prospective study and systematic review. PLoS Med 4, e270, doi:10.1371/journal.pmed.0040270 (2007).

7 Masson, P. et al. Chronic kidney disease and the risk of stroke: a systematic review and meta-analysis. Nephrol Dial Transplant 30, 1162–1169, doi:10.1093/ndt/gfv009 (2015).

8 Chronic Kidney Disease Prognosis, C. et al. Association of estimated glomerular filtration rate and albuminuria with all-cause and cardiovascular mortality in general population cohorts: a collaborative meta-analysis. Lancet 375, 2073-2081, doi:10.1016/S0140-6736(10)60674-5 (2010).

9 Major, R. W. et al. Cardiovascular disease risk factors in chronic kidney disease: A systematic review and meta-analysis. PloS one 13, e0192895, doi:10.1371/journal.pone.0192895 (2018).

10 Aragam, K. G. et al. Discovery and systematic characterization of risk variants and genes for coronary artery disease in over a million participants. Nat Genet 54, 1803–1815, doi:10.1038/s41588-022-01233-6 (2022).

11 Tcheandjieu, C. et al. Large-scale genome-wide association study of coronary artery disease in genetically diverse populations. Nat Med 28, 1679–1692, doi:10.1038/s41591-022-01891-3 (2022).

12 Malik, R. et al. Multiancestry genome-wide association study of 520,000 subjects identifies 32 loci associated with stroke and stroke subtypes. Nat Genet 50, 524–537, doi:10.1038/s41588-018-0058-3 (2018).

13 Chen, J. et al. Coronary Artery Calcification and Risk of Cardiovascular Disease and Death Among Patients With Chronic Kidney Disease. JAMA Cardiol 2, 635–643, doi:10.1001/jamacardio.2017.0363 (2017).

14 Denker, M. et al. Chronic Renal Insufficiency Cohort Study (CRIC): Overview and Summary of Selected Findings. Clin J Am Soc Nephrol 10, 2073–2083, doi:10.2215/CJN.04260415 (2015).

15 Mishra, A. et al. Stroke genetics informs drug discovery and risk prediction across ancestries. Nature 611, 115–123, doi:10.1038/s41586-022-05165-3 (2022).

16 Ibanez, L. et al. Multi-ancestry GWAS reveals excitotoxicity associated with outcome after ischaemic stroke. Brain 145, 2394–2406, doi:10.1093/brain/awac080 (2022).

17 Lash, J. P. et al. Chronic Renal Insufficiency Cohort (CRIC) Study: baseline characteristics and associations with kidney function. Clin J Am Soc Nephrol 4, 1302–1311, doi:10.2215/CJN.00070109 (2009).

18 Feldman, H. I. et al. The Chronic Renal Insufficiency Cohort (CRIC) Study: Design and Methods. J Am Soc Nephrol 14, S148–153, doi:10.1097/01.asn.0000070149.78399.ce (2003).

19 Inker, L. A. et al. New Creatinine- and Cystatin C-Based Equations to Estimate GFR without Race. The New England journal of medicine 385, 1737–1749, doi:10.1056/NEJMoa2102953 (2021).

20 Parsa, A. et al. Genome-Wide Association of CKD Progression: The Chronic Renal Insufficiency Cohort Study. J Am Soc Nephrol 28, 923–934, doi:10.1681/ASN.2015101152 (2017).

21 Fang, H. et al. Harmonizing Genetic Ancestry and Self-identified Race/Ethnicity in Genome-wide Association Studies. American journal of human genetics 105, 763–772, doi:10.1016/j.ajhg.2019.08.012 (2019).

22 Price, A. L. et al. Principal components analysis corrects for stratification in genome-wide association studies. Nat Genet 38, 904–909, doi:10.1038/ng1847 (2006).

23 Fuchsberger, C., Abecasis, G. R. & Hinds, D. A. minimac2: faster genotype imputation. Bioinformatics 31, 782–784, doi:10.1093/bioinformatics/btu704 (2015).

24 Chen, H. et al. Control for Population Structure and Relatedness for Binary Traits in Genetic Association Studies via Logistic Mixed Models. American journal of human genetics 98, 653–666, doi:10.1016/j.ajhg.2016.02.012 (2016).

25 Willer, C. J., Li, Y. & Abecasis, G. R. METAL: fast and efficient meta-analysis of genomewide association scans. Bioinformatics 26, 2190–2191, doi:10.1093/bioinformatics/btq340 (2010).

26 Huang, L. et al. TOP-LD: A tool to explore linkage disequilibrium with TOPMed whole-genome sequence data. American journal of human genetics 109, 1175–1181, doi:10.1016/j.ajhg.2022.04.006 (2022).

27 Sudlow, C. et al. UK biobank: an open access resource for identifying the causes of a wide range of complex diseases of middle and old age. PLoS Med 12, e1001779, doi:10.1371/journal.pmed.1001779 (2015).

28 Wen, J. et al. Transcriptome-Wide Association Study of Blood Cell Traits in African Ancestry and Hispanic/Latino Populations. Genes (Basel) 12, doi:10.3390/genes12071049 (2021).

29 Atkinson, E. G. et al. Tractor uses local ancestry to enable the inclusion of admixed individuals in GWAS and to boost power. Nat Genet 53, 195–204, doi:10.1038/s41588-020-00766-y (2021).

30 Browning, S. R. et al. Local Ancestry Inference in a Large US-Based Hispanic/Latino Study: Hispanic Community Health Study/Study of Latinos (HCHS/SOL). G3 6, 1525–1534, doi:10.1534/g3.116.028779 (2016).

31 Mbatchou, J. et al. Computationally efficient whole-genome regression for quantitative and binary traits. Nat Genet 53, 1097–1103, doi:10.1038/s41588-021-00870-7 (2021).

32 Bulik-Sullivan, B. K. et al. LD Score regression distinguishes confounding from polygenicity in genome-wide association studies. Nat Genet 47, 291–295, doi:10.1038/ng.3211 (2015).

