## Supplementary materials for "Genetics of cardiovascular outcomes in individuals with chronic kidney disease: the Chronic Renal Insufficiency Cohort (CRIC) study"

*Shared authors

**Affiliations**

1 Department of Genetics, University of North Carolina, Chapel Hill, NC, USA
2 Department of Biostatistics, University of North Carolina, Chapel Hill, NC, USA
3 Department of Applied Physical Sciences, University of North Carolina, Chapel Hill, NC, USA
4 Department of Epidemiology, Rollins School of Public Health, Emory University, Atlanta, Georgia, USA
5. Veterans Affairs Atlanta Healthcare System, Decatur, Georgia, USA
6. Department of Statistics and Operations Research, University of North Carolina at Chapel Hill, Chapel Hill, NC, USA
7. Division of Research, Kaiser Permanente Northern California, Oakland, California, USA
8. Departments of Epidemiology, Biostatistics and Medicine, University of California, San Francisco, San Francisco, California; Department of Medicine (Nephrology), Stanford University, Palo Alto, California, USA
9. Division of Genetic Medicine, Vanderbilt University Medical Center, Nashville, TN, USA
10. Vanderbilt Genetics Institute, Vanderbilt University Medical Center, Nashville, TN, USA
11. Department of Medicine, University of Pennsylvania, Philadelphia, Pennsylvania, USA
12. Department of Medicine (Nephrology), Case Western Reserve University/University Hospitals Cleveland Medical Center, Cleveland, OH, USA
13. Division of Nephrology and Hypertension, University Hospitals Cleveland Medical Center, Louis Stokes Cleveland Veterans Affairs Medical Center, Case Western Reserve University, Cleveland, Ohio, USA
14. Department of Medicine, University of Illinois, Chicago, Illinois, USA
15. Massachusetts Veterans Epidemiology Research and Information Center (MAVERIC), VA Boston Healthcare System, Boston, MA, USA.
16. VA Tennessee Valley Healthcare System, Nashville, TN, USA.
17. Division of Nephrology and Hypertension, Vanderbilt Center for Kidney Disease, Department of Medicine, Vanderbilt University Medical Center, Nashville, TN, USA
18. Department of Medicine, Brigham & Women's Hospital and Harvard Medical School, Boston, Massachusetts, USA
19. Division of Cardiology, Emory University School of Medicine, Atlanta, Georgia, USA.
20. Department of Epidemiology, University of North Carolina, Chapel Hill, NC, USA

§ Lawrence J. Appel, MD, MPH, Jing Chen, MD, MMSc, MSc, Debbie L Cohen, MD, Harold I. Feldman, MD, MSCE, Alan S. Go, MD, James P. Lash, MD, Robert G. Nelson, MD, PhD, MS, Mahboob Rahman, MD, Panduranga S. Rao, MD, Vallabh O Shah, PhD, MS, Mark L. Unruh, MD, MS

Corresponding authors:
Nora Franceschini, MD, MPH, Departments of Epidemiology and Biostatistics
Yun Li, PhD, University of North Carolina, Chapel Hill, NC

### Supplementary Information

#### Time-to-event analysis

#### Method

In the time-to-event analysis, we obtained martingale residuals from Cox proportional hazard models adjusted for covariates, and martingale residuals were regressed into SNVs in mixed linear regression models implemented by EPACTS v3.3.0. The results were then combined using fixed-effect meta-analysis[1]. The significance threshold was $5x10^{-8}$ to account for SNPs tested and traits. We attempted replication in the UK Biobank using events identified after the baseline assessments and logistic models adjusted for the same covariates described in the Replication section.

#### Results

**Time-to event results.** Findings for analyses restricted to incident events in time-to-event analysis are shown in **Table S2 and Table S3.** There was little evidence for genome inflation for incident CHD ($\lambda$ =1.0073), incident non-fatal MI ($\lambda$ =1.0121), incident all-cause stroke ($\lambda$ = 1.0128), and incident ischemic stroke ($\lambda$ =1.0145). Meta-analysis of AFR and EUR CRIC samples identified two common intronic variants with MAF > 3% at *SEMA5A* (incident fatal/non-fatal CHD), two common intronic variants with MAF > 3% at *ABCB5* (incident ischemic stroke) and only SNV at *ABCB5* (incident all-cause stroke) at *p*.value < 5e-8 (**Table S3**).

**UK Biobank replication.** We attempted to replicate 5 variants spanning 2 loci in UK Biobank replication (**Table S3**), and two variants at *ABCB5* were associated with incident ischemic stroke at the nominal *p*.value < 0.05 (**Table S3**). These two variants had the same direction of association between discovery and replication (**Table 2 and Table S3, Supplementary materials)**.

**Comparison of our findings with published GWAS of CHD and stroke outcomes.** When comparing the time-to-event analysis results with published GWAS, there were in total 5 variants residing in 3 published loci (any variant within the loci < 1MB from the CRIC index variant) across four incident events in time-to-event analysis (**Table S7 and S8**) but can't find any LD between the variants discovered in time-to-event analysis and the published variants associated with CHD or any type of stroke**.**


#### Supplementary Acknowledgements

**VA Million Veteran Program:**
**Core Acknowledgements for Publications**
**May 2024**

#### MVP Program Office

- Sumitra Muralidhar, Ph.D., Program Director
  US Department of Veterans Affairs, 810 Vermont Avenue NW, Washington, DC 20420
- Jennifer Moser, Ph.D., Associate Director, Scientific Programs
  US Department of Veterans Affairs, 810 Vermont Avenue NW, Washington, DC 20420
- Jennifer E. Deen, B.S., Associate Director, Cohort & Public Relations
  US Department of Veterans Affairs, 810 Vermont Avenue NW, Washington, DC 20420

**MVP Executive Committee**

- Co-Chair: Philip S. Tsao, Ph.D.
  VA Palo Alto Health Care System, 3801 Miranda Avenue, Palo Alto, CA 94304
- Co-Chair: Sumitra Muralidhar, Ph.D.
  US Department of Veterans Affairs, 810 Vermont Avenue NW, Washington, DC 20420
- J. Michael Gaziano, M.D., M.P.H.
  VA Boston Healthcare System, 150 S. Huntington Avenue, Boston, MA 02130
- Elizabeth Hauser, Ph.D.
  Durham VA Medical Center, 508 Fulton Street, Durham, NC 27705
- Amy Kilbourne, Ph.D., M.P.H.
  VA HSR&D, 2215 Fuller Road, Ann Arbor, MI 48105
- Michael Matheny, M.D., M.S., M.P.H.
  VA Tennessee Valley Healthcare System, 1310 24th Ave. South, Nashville, TN 37212
- Dave Oslin, M.D.
  Philadelphia VA Medical Center, 3900 Woodland Avenue, Philadelphia, PA 19104
- Deepak Voora, MD
  Durham VA Medical Center, 508 Fulton Street, Durham, NC 27705

**MVP Co-Principal Investigators**

- J. Michael Gaziano, M.D., M.P.H.
  VA Boston Healthcare System, 150 S. Huntington Avenue, Boston, MA 02130
- Philip S. Tsao, Ph.D.
  VA Palo Alto Health Care System, 3801 Miranda Avenue, Palo Alto, CA 94304

**MVP Core Operations**

- Jessica V. Brewer, M.P.H., Director, MVP Cohort Operations
  VA Boston Healthcare System, 150 S. Huntington Avenue, Boston, MA 02130
- Mary T. Brophy M.D., M.P.H., Director, VA Central Biorepository
  VA Boston Healthcare System, 150 S. Huntington Avenue, Boston, MA 02130
- Kelly Cho, M.P.H, Ph.D., Director, MVP Phenomics
  VA Boston Healthcare System, 150 S. Huntington Avenue, Boston, MA 02130
- Lori Churby, B.S., Director, MVP Regulatory Affairs
  VA Palo Alto Health Care System, 3801 Miranda Avenue, Palo Alto, CA 94304
- Scott L. DuVall, Ph.D., Director, VA Informatics and Computing Infrastructure (VINCI)
  VA Salt Lake City Health Care System, 500 Foothill Drive, Salt Lake City, UT 84148
- Saiju Pyarajan Ph.D., Director, Data and Computational Sciences
  VA Boston Healthcare System, 150 S. Huntington Avenue, Boston, MA 02130
- Robert Ringer, Pharm.D., Director, VA Albuquerque Central Biorepository
  New Mexico VA Health Care System, 1501 San Pedro Drive SE, Albuquerque, NM 87108
- Luis E. Selva, Ph.D., Director, MVP Biorepository Coordination
  VA Boston Healthcare System, 150 S. Huntington Avenue, Boston, MA 02130
- Shahpoor (Alex) Shayan, M.S., Director, MVP PRE Informatics

VA Boston Healthcare System, 150 S. Huntington Avenue, Boston, MA 02130

- Brady Stephens, M.S., Principal Investigator, MVP Information Center
  Canandaigua VA Medical Center, 400 Fort Hill Avenue, Canandaigua, NY 14424
- Stacey B. Whitbourne, Ph.D., Director, MVP Cohort Development and Management
  VA Boston Healthcare System, 150 S. Huntington Avenue, Boston, MA 02130

**MVP Publications and Presentations Committee**

- Co-Chair: Themistocles L. Assimes, M.D., Ph. D
  VA Palo Alto Health Care System, 3801 Miranda Avenue, Palo Alto, CA 94304
- Co-Chair: Adriana Hung, M.D.; M.P.H
  VA Tennessee Valley Healthcare System, 1310 24th Ave. South, Nashville, TN 37212
- Co-Chair: Henry Kranzler, M.D.
  Philadelphia VA Medical Center, 3900 Woodland Avenue, Philadelphia, PA 19104

**Supplementary Tables and Figures**

**Table S1**. Cardiovascular disease events used in CRIC.

**Table S2**. Significant GWAS results for four incident and two combined incident/prevalent outcomes tested. The lowest *p*.value SNV for each locus are reported, and the MAF is the average of between AFR and EUR MAFs.

**Table S3**. Significant GWAS AFR/EUR CRIC meta-analysis results for incident outcomes.

**Table S4**. Genetic correlations among MVP, published GWAS and CRIC for cardiovascular outcomes.

**Table S5**. Published variants tested in the CRIC study.

**Table S6.** Characteristics of published GWAS meta-analysis for CHD and Stroke that were used to compare associations in CRIC.

**Table S7**. Association results from published SNVs in CRIC AFR+EUR for outcomes.

**Table S8**. Association results from published SNVs in CRIC EUR.

**Table S9**. The number replicated loci of MVP for the published known GWAS.

**Figure S1**. **A**. Manhattan plots for incident CHD in CRIC. X-axis lists the chromosome position for each variant and Y-axis shows the -log10(*p*.value) for associations. **B**. Quantile-quantile (QQ) plot of GWAS results for incident CHD in CRIC

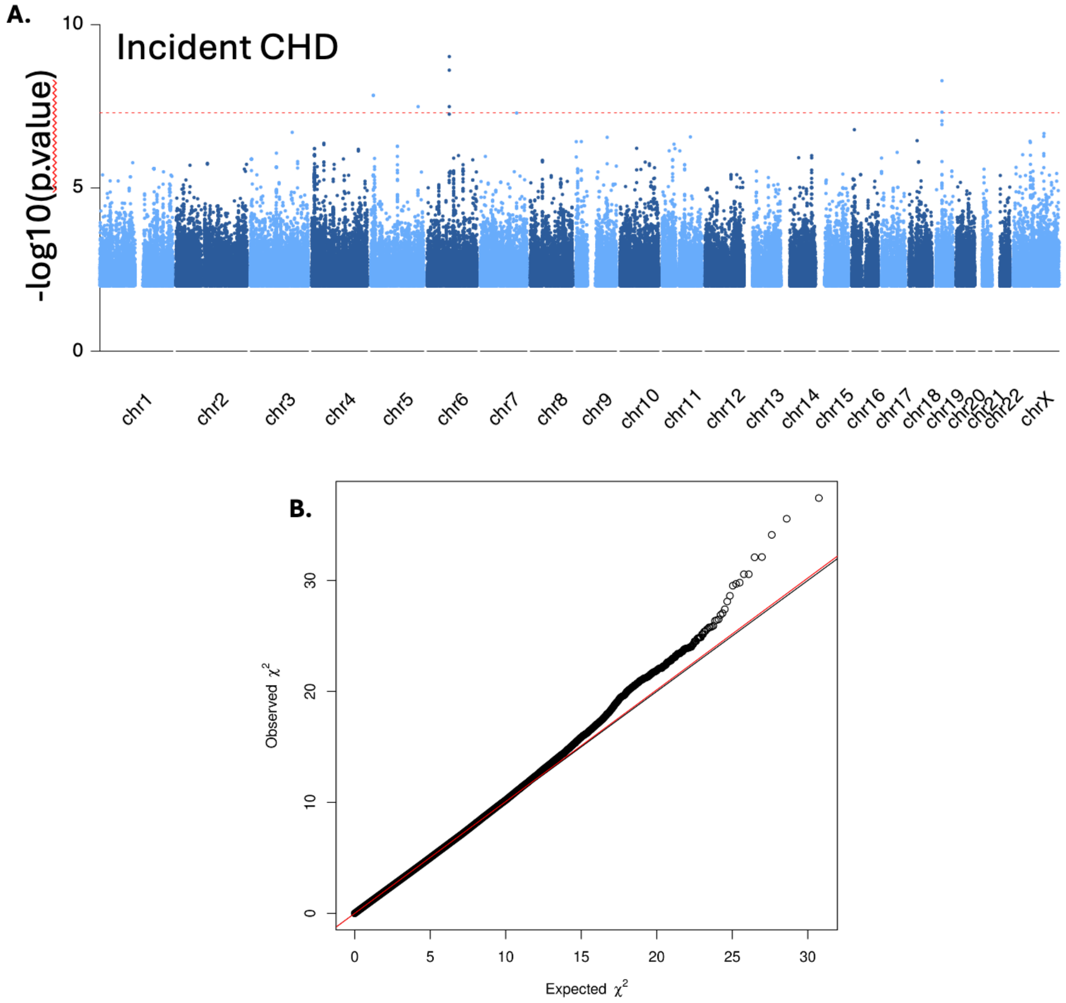

**Figure S2**. **A**. Manhattan plots for incident non-fatal CHD in CRIC. X-axis lists the chromosome position for each variant and Y-axis shows the -log10(*p*.value) for associations. **B**. Quantile-quantile (QQ) plot of GWAS results for incident non-fatal CHD in CRIC

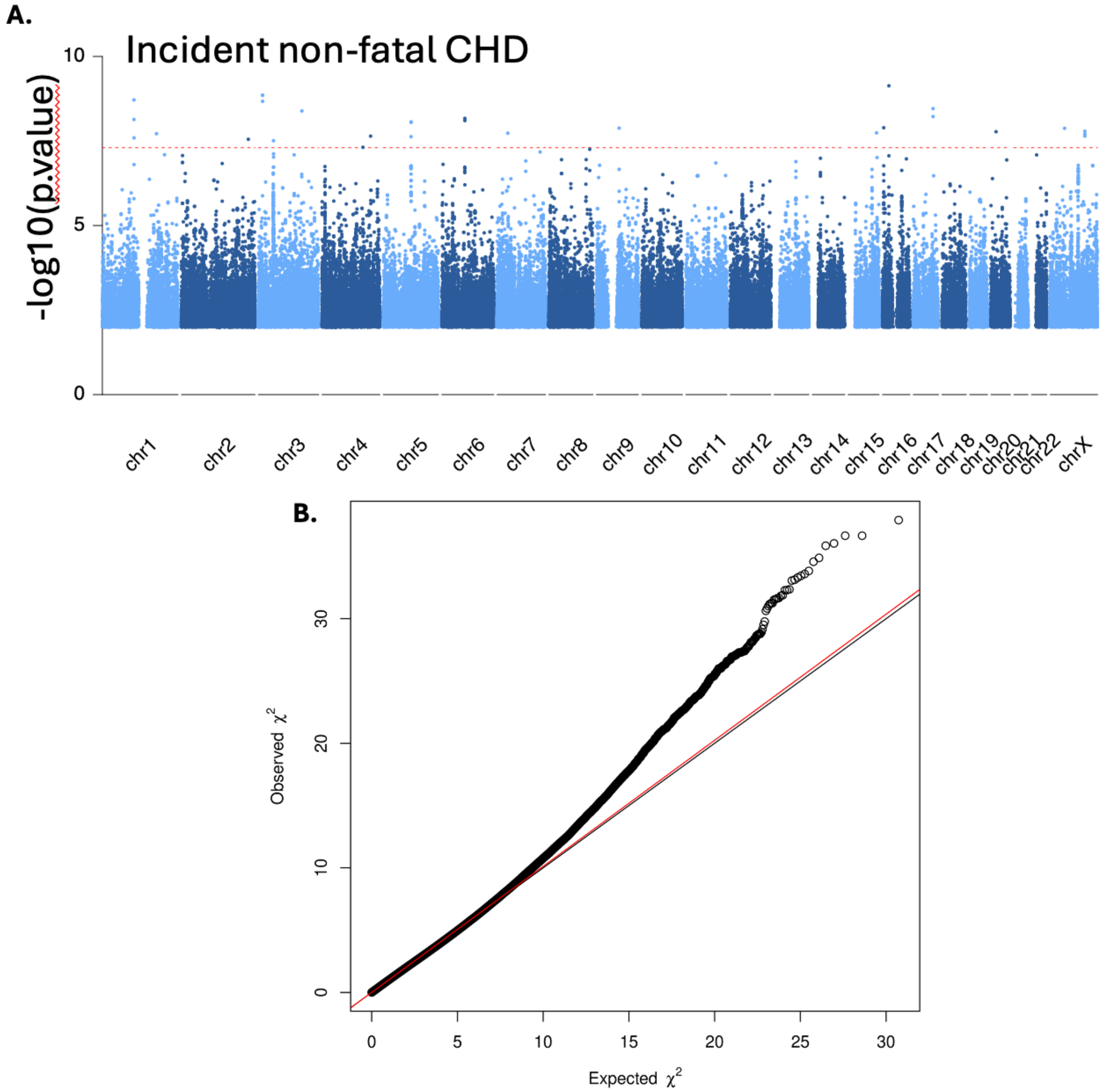

**Figure S3**. **A**. Manhattan plots for incident all-cause stroke in CRIC. X-axis lists the chromosome position for each variant and Y-axis shows the -log10(*p*.value) for associations. **B**. Quantile-quantile (QQ) plot of GWAS results for incident all-cause stroke in CRIC

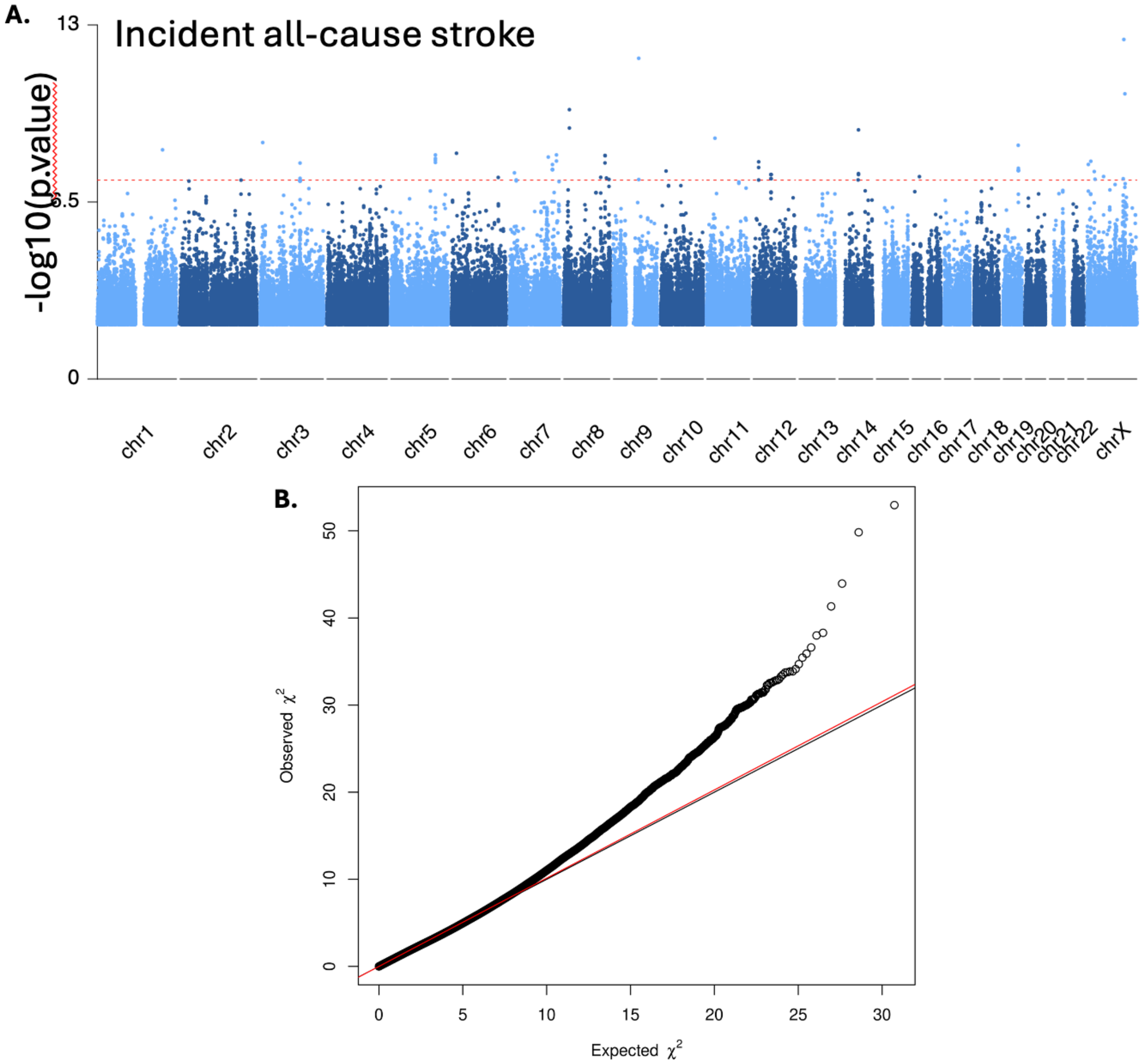

**Figure S4**. A. Manhattan plots for incident ischemic stroke in CRIC. X-axis lists the chromosome position for each variant and Y-axis shows the -log10(*p*.value) for associations. B. Quantile-quantile (QQ) plot of GWAS results for incident ischemic stroke in CRIC.

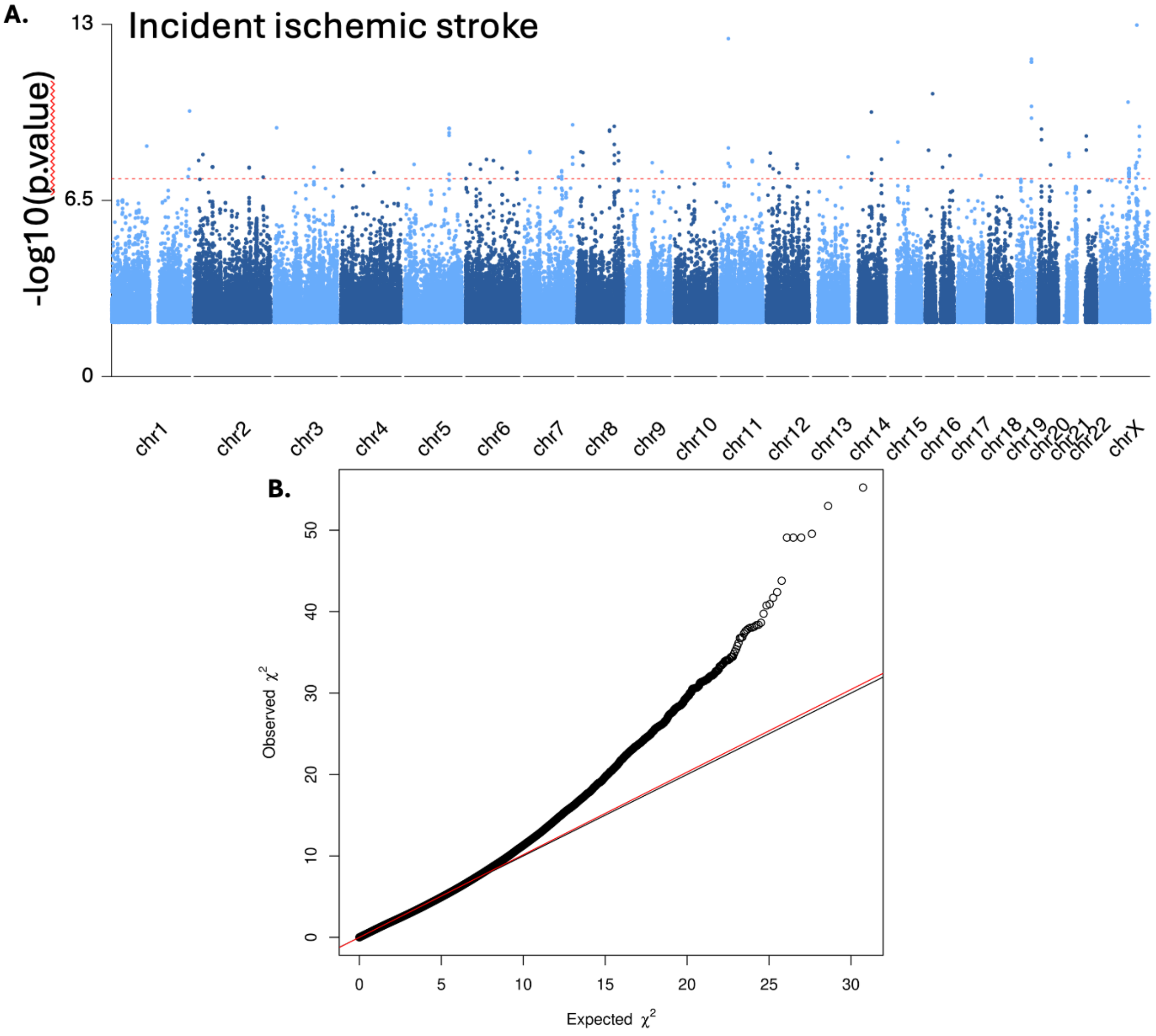

### Reference

1 Willer, C. J., Li, Y. & Abecasis, G. R. METAL: fast and efficient meta-analysis of genomewide association scans. *Bioinformatics* **26**, 2190-2191, doi:10.1093/bioinformatics/btq340 (2010).
